# Can We Really Trust the Findings of the COVID-19 Research? Quality Assessment of Randomized Controlled Trials Published on COVID-19

**DOI:** 10.1101/2022.04.15.22273881

**Authors:** Athira S Joshy, Christy Thomas, Saphal Surendran, Krishna Undela

## Abstract

**Objective:** To evaluate the quality of randomized controlled trials (RCTs) published on Coronavirus Disease-19 (COVID-19) and to investigate the reasons behind compromising the quality, if found.

**Methods:** A systematic literature search was performed in PubMed, Google Scholar, and Cochrane CENTRAL to identify the Randomized Controlled Trails published on Coronavirus Disease-19 between 1^st^ Dec 2019 to 31^st^ Aug 2021. Research articles met with study criteria were included in the study. Assessment of quality of randomized controlled trials was done using modified Jadad scale.

**Results:** 21,259 records of randomized controlled trials were identified through database searching, out of which 90 randomized controlled trials were included in the study and, 34 (37.8%) were of high-quality, 46 (51.1%) were of moderate quality, and 10 (11.1 %) were of low-quality studies. There were 40 (44.4%), 38 (42.2%), and 12 (13.3%) randomized controlled trials published in the early, middle, and late terms with Jadad score 5.12±1.67, 5.34±1.32, and 5.68±1.50 respectively (P=0.52). When comparing the blinding status, appropriate blinding, and methods to evaluate adverse events in randomized controlled trials with modified Jadad score, a significant difference was observed (P<0.001). A significant moderate positive correlation was found between the impact factor of the journal and the modified Jadad scale score (R2= 0.48, P<0.001).

**Conclusion:** Findings from our study indicate that accelerated publication of Coronavirus Disease-19 researches along with the fast-track review process has resulted in lowering study quality scores. With the emergence of stronger evidence, Coronavirus Disease-19 clinical studies with lower methodological quality should be revisited.

**Impacts on practice:**

- There have been numerous sacrifices and tragedies in the clinical response to covid-19. Revising the quality of randomized controlled trials published on COVID-19 as we enter the third wave of the pandemic and beyond, will improve the evidence-based practice of medications for clinical pharmacy services.
- COVID-19 Patients will benefit from evidence-based pharmaceutical care through reduced drug-related problems.

## 1. INTRODUCTION

Coronavirus disease 2019 (COVID-19), was first reported on 31^st^ December 2019 in Wuhan City, Hubei Province, China [1]. SARS-CoV-2 (Severe Acute Respiratory Syndrome Coronavirus) is the disease-causing agent of COVID-19. Concerned about the disease’s rapid spread, the World Health Organization (WHO) declared it a pandemic on March 11, 2020. As of January 27, 2022, WHO had confirmed 364,191,494 COVID-19 cases, with 5,631,457 deaths [2], and Omicron, a novel SARS-CoV-2 variation of concern, has been reported [3].

Since January 2020, the scientific community has responded enthusiastically to the COVID-19, with information being released at a breakneck pace. As of the time of writing, ClinicalTrials.gov had 7,047 trials registered while WHO’s International Clinical Trial Registry Platform has 12,508 trials related to COVID-19 [4]. Rapid progression of the COVID-19 pandemic combined with a lack of available knowledge to guide clinical decision-making is the reason for this exponential increase in the publication of research articles on COVID-19, in the past year [5]. WHO is bringing together scientists and global health professionals from all over the world to speed up research and development, as well as set new norms and standards to manage the coronavirus pandemic and to give proper care to the infected individuals. However, the reliability of the findings of these studies depends on the quality of the research.

The randomized controlled trial (RCT) is the recognized gold standard in clinical research and is thought to produce the best level of evidence for clinically assessing competing interventions, followed by clinical trials. RCT findings are used in Evidence-Based Medicine (EBM) because they are built on an objective, scientific, and systematic study design [6]. The peer-review and fast-track publication process may have inadvertent effects, leading to flawed methodology, and poor quality of findings on COVID-19 [7]. The COVID-19 pandemic’s urgency and severity pose both challenges and possibilities for practitioners interested in using EBM [8] and indeed the low quality of published RCTs are thrones in the road of EBM practice.

In a methodological review on the quality of COVID-19 research, authors evaluated the quality of observational studies (cohort and case-control studies), descriptive studies (case-series), and RCTs published in selected journals [9]. A study reported by Jung et.al. concluded that there were methodological and reporting issues in the articles published on COVID-19 in major clinical journals, that can result in compromising the utility of the research [10]. Both studies, however, failed to include research articles published in all relevant journals or to focus more on RCTs, which are the highest level of evidence in the evidence-based hierarchy. To the best of our knowledge, there are no studies conducted to date to find out the quality of RCTs research articles published on COVID-19. Hence, this study aimed to evaluate the quality of RCTs to investigate the reasons behind compromising the quality.

## 2. METHODOLOGY

This is a retrospective quality assessment study. Research articles were included in the study based on the following inclusion and exclusion criteria. Full-text articles of RCTs conducted either for the prevention, diagnosis, management, and treatment of COVID-19 and which were published in the English language from 1^st^ December 2019 to 31^st^ August 2021were included in the study. All the *in-vitro* or *in-vivo* studies, descriptive studies (Case series, Case reports, Ecological studies, Descriptive cross-sessional studies), observational cross-sessional studies, non-randomized controlled trials, studies unavailable in the electronic form and conference abstract, and some other studies (Study protocols, post hoc analyses, brief reports, study design & rationale, letters, and correspondence) were excluded from our study.

### 2.1. Search strategy

The electronic databases such as PubMed, Google Scholar, and Cochrane CENTRAL were searched from 1^st^ December 2019 to 31^st^ August 2021 MeSH (Medical Subject Headings) terms were identified and were combined using Boolean operators while searching in PubMed. The search strategy followed in PubMed for RCTs was ((“covid 19”[Supplementary Concept] OR “Coronavirus Infections”[MeSH Terms:noexp] OR “severe acute respiratory syndrome coronavirus 2”[Supplementary Concept]) AND (“randomized controlled trial”[Publication Type] OR (“controlled clinical trial”[Publication Type])). By using the same keywords, a search was also performed in Google Scholar and Cochrane Central, for identifying the articles published in journals that were not indexed in PubMed. The detailed search strategy can be found in the supplementary appendix (S1).

### 2.2. Data collection and analysis

#### 2.2.1. Study selection

The author (AJ) independently screened the titles and abstracts of the search results and will code them as either ‘retrieve’ (eligible or potentially eligible/unclear) or ‘do not retrieve’. We retrieved the full[text study reports of all potentially eligible studies. Two review authors (AJ and SS) independently assessed these for inclusion and recorded the reasons for the exclusion of ineligible studies. We resolved any disagreement through discussion or, if required, we consulted a third review author (KU).

#### 2.2.2. Data extraction and management

Research articles that met the study criteria were included in the study and data extraction was performed using Microsoft Excel. The following study characteristics were extracted from the included studies (if applicable); the title of the article, name of the journal, name of the authors, name of the corresponding author, name of the corresponding author’s institution, name of the publisher, country in which the study was conducted, type of the study design, year of publication, volume number, issue number, page number of the article and the information required for quality assessment of the articles.

### 2.3. Assessment of quality of the study

The quality of RCTs was assessed using the Modified Jadad scale (also known as the Oxford quality scoring system). Using this scale, studies were scored according to the presence of six features of RCTs, like randomization, blinding, withdrawals or dropouts, inclusion or exclusion criteria, statistical analysis, and the method used to assess the adverse events. Scale scores ranging from zero to eight [11,12] Two reviewers (AJ and SS) have independently assessed the quality of included studies using the Modified Jadad Scale (Supplementary appendix S2). Disagreements between the authors were resolved by consensus with a third reviewer (KU). Based on scores obtained from the scale, studies were categorized into high-quality (with a score of 6-8 points), moderate quality (4-5 points), or poor-quality (<4 points) out of a total score of 8.

### 2.4. Statistical analysis

The studies were divided into three terms: Early (2019 December to 2020 July), mid (2020 August to 2021 January), and late (2021 February to 2021 August). Counted data are described by a number and percentage. The scores obtained by Jadad score were compared and analyzed using one-way analysis of variance test whereas different quality article ratio was analyzed by chi-square test. In addition, the comparisons of different factors influencing the quality of the studies were analyzed using the Student t-test. SPSS v.22.0 was used for all statistical analysis and a P-value of <0.05 was considered statistically significant.

## 3. RESULTS

From a total of 21,259 studies identified through different databases (PubMed=11,763, Google Scholar=8,128, Cochrane CENTRAL=1,368), 90 RCTs met with study criteria were included in the study [13–102](Figure:1). Out of 90 RCTs, 59 (65.6%) were conducted for treatment, 21 (23.3%) were conducted for prevention, seven (7.8%) were conducted for management, three (3.3%) were conducted for the diagnosis of COVID-19 (Figure 2). A total of 55 (61.1%) RCTs included in the study were found to be open-labeled studies and only 26 (28.9%) studies were double-blinded (Figure 3).

**Figure 1:**
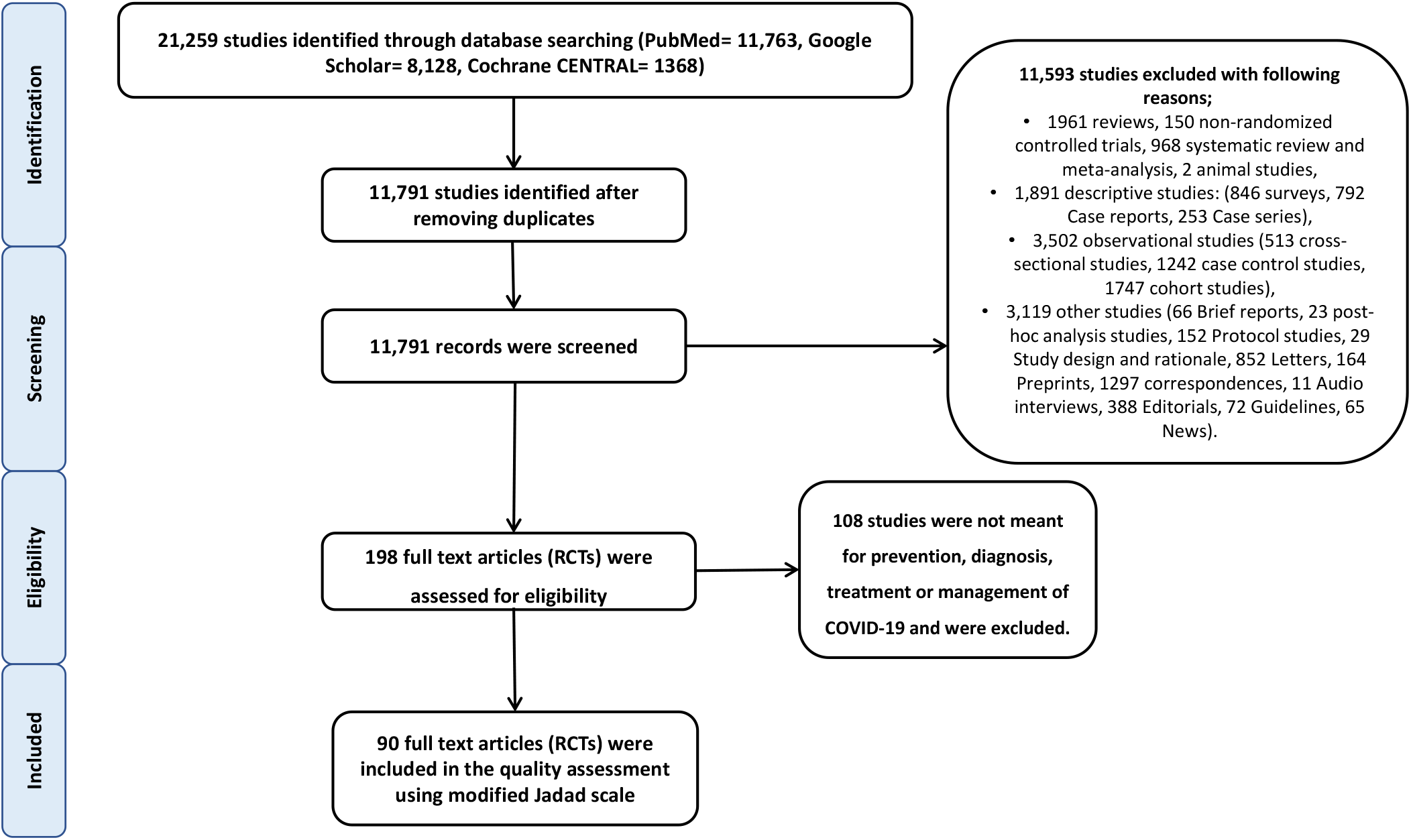
Study flow diagram of the review selection.

**Figure 2:**
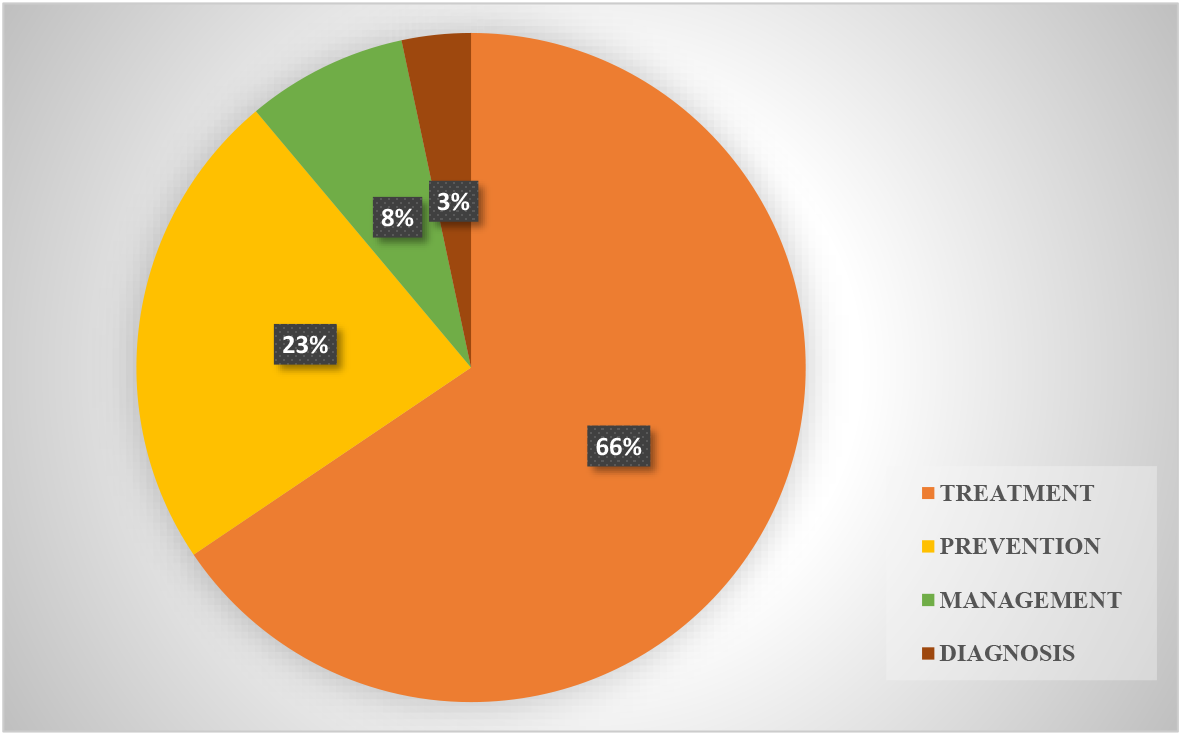
The 90 RCTs with the reason for conducting the trial.

**Figure 3:**
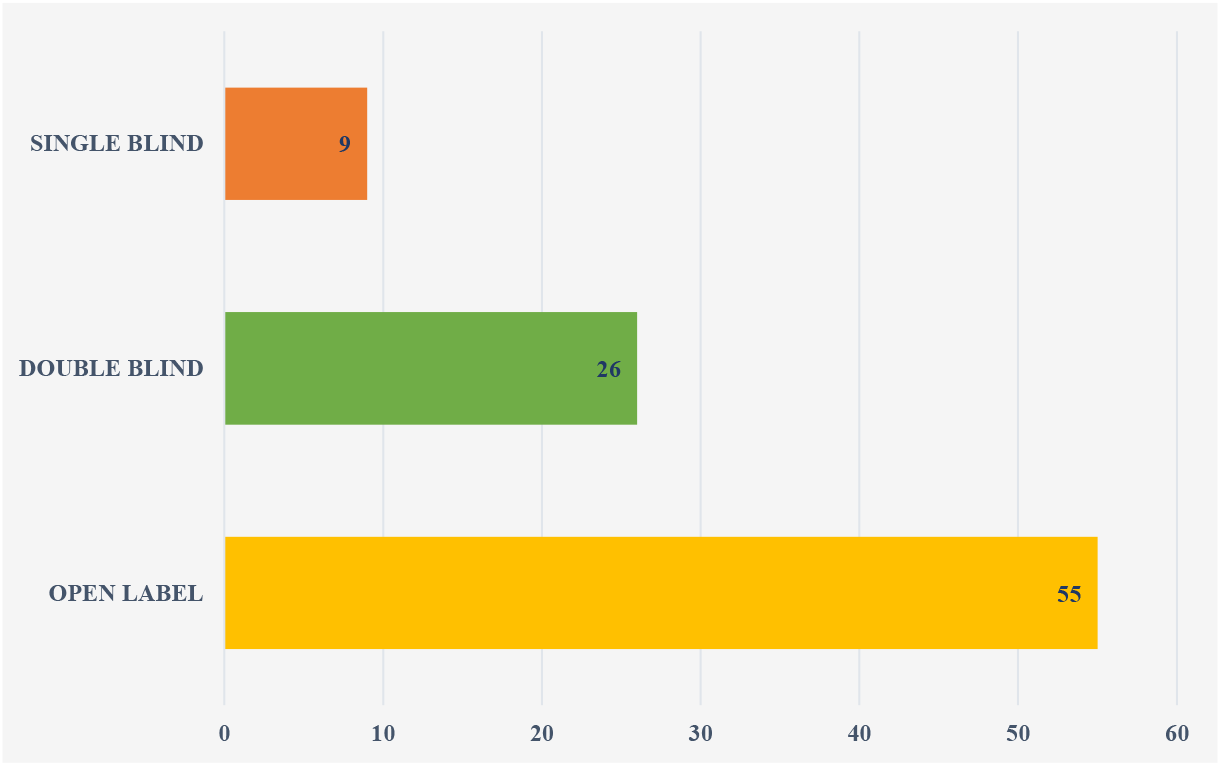
The 92 RCTs with the type of blinding.

Out of 90 RCTs, 34 (37.8%) were of high-quality, 46 (51.1%) were of moderate quality, and 10 (11.1 %) were of low-quality studies (Table 1). According to the Jadad scale, 26 RCTs (28.9%) reported blinding, whereas only 22 (24.4%) reported adequate blinding. Only 36 (40.0%) of the 90 RCTs were reported with methodologies to assess adverse events (Table 2).

**Table 1:**
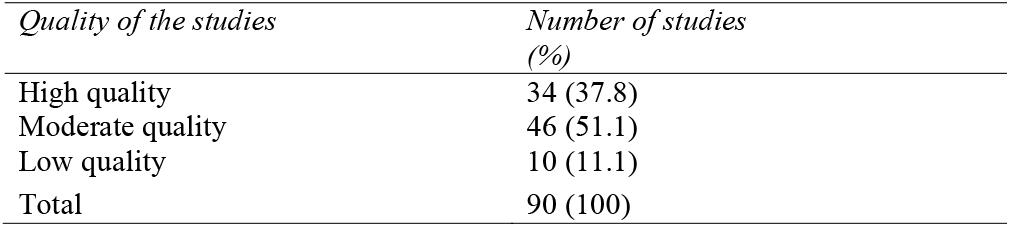
Quality of the studies.

| *Quality of the studies* | *Number of studies (%)* |
| --- | --- |
| High quality | 34 (37.8) |
| Moderate quality | 46 (51.1) |
| Low quality | 10 (11.1) |
| Total | 90 (100) |

**Table 2:** Components of Jaded scale and number of studies.

| *Components of the scale* | *Number of RCTs (%)* |  |
| --- | --- | --- |
|  | *Yes* | *No* |
| Was the study described as Randomised? | 90 (100) | 0 (0) |
| Was the method of randomization appropriate? | 68 (75.6) | 22 (24.4) |
| Double blinding is reported | 26 (28.9) | 64 (71.1) |
| Double blinding is appropriate | 22 (24.4) | 68 (75.6) |
| Withdrawals are reported as number and reason per arm | 60 (66.7) | 30 (33.3) |
| Method(s) used to assess adverse events is described | 36 (40.0) | 54 (60.0) |
| Methods of statistical analysis are described | 81 (90.0) | 9 (10.0) |
| Inclusion and/or exclusion criteria are reported | 87 (96.7) | 3 (3.3) |

RCT: Randomised Controlled trial.

### 3.1. Qualitative variations of RCTs by the term

There were 40 (44.4%), 38 (42.2%), and 12 (13.3%) papers published in the early, middle, and late terms, respectively. The Jadad score for the early, mid, and late terms were 5.12±1.67, 5.34±1.32, and 5.68±1.50 respectively (P=0.52). The number of high-quality articles for the early, mid, and late terms is 12 (30.0), 14(36.8), and 8(66.7) respectively. There was no significant difference observed between the number of different quality studies and different publication terms (P=0.097, Table 3).

**Table 3:**
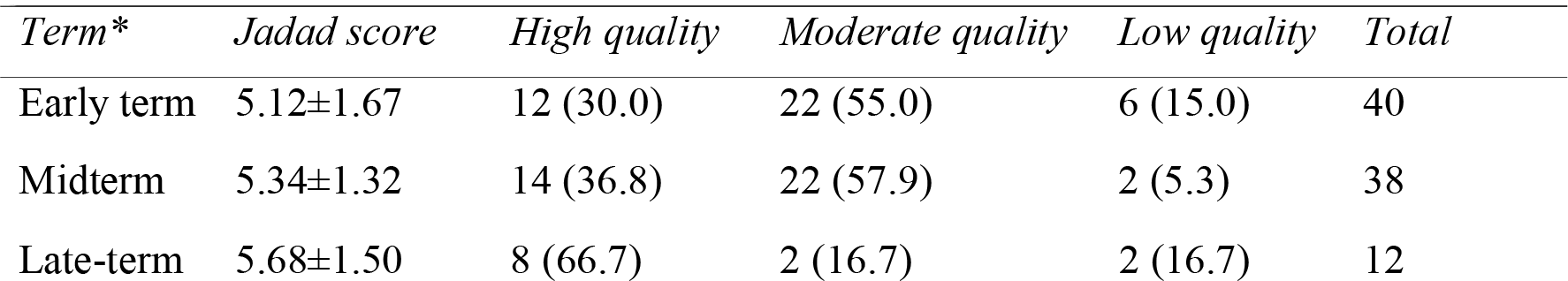
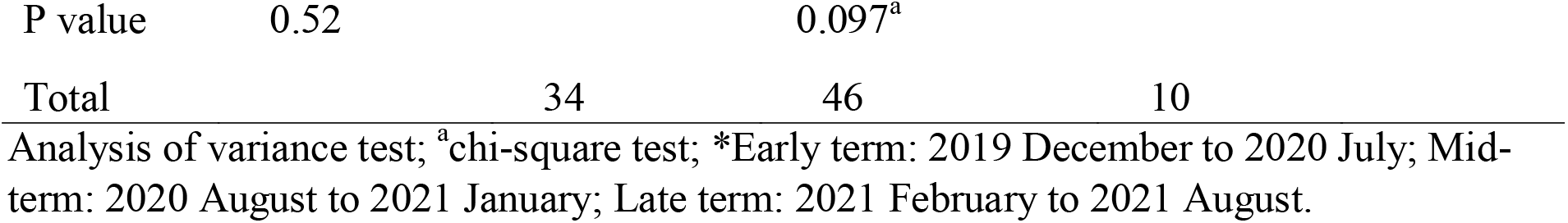
Quality assessment of randomized controlled trials according to the publication term.

### 3.2. Analysis of factors related to the quality of the articles

It was noted that there was a statistically significant difference in the mean modified jaded scale score and number of studies by blinding status in the trials (P<0.001). Similarly, in trials with and without appropriate blinding, the mean modified Jadad scale was 7.25±0.80 and 4.59±1.01, respectively, with the difference in the score is statistically significant (P<0.001). There was also a significant variation in the number of RCTs in different quality levels and appropriate blinding (P<0.001). The mean modified Jadad scale score in RCTs mentioned with and without methods to assess adverse events were 6.12±1.43 and 4.65±1.22 respectively (P<0.001; Table 4). There was a positive correlation noticed between the impact factor of the journal and the modified Jadad scale score (R2= 0.48), which was significant at the level of P<0.001.

**Table 4:** Factors associated with the quality of Randomized Controlled Trials.

| *factors* | *No. of RCTs (%)* | *Jadad Score* | *High quality* | *Moderate quality* | *Low quality* |
| --- | --- | --- | --- | --- | --- |
| Blinding |  |  |  |  |  |
| Yes | 26 (28.9) | 7.00±1.01 | 23 (25.6) | 3 (3.3) | 0 (0) |
| No | 64 (71.1) | 4.53±0.98 | 11 (12.2) | 43 (47.8) | 10 (11.1) |
| P value |  | <0.001 | <0.001<sup>a</sup> |  |  |
| Appropriate Blinding |  |  |  |  |  |
| Yes | 22 (24.4) | 7.25±0.80 | 21 (23.3) | 1 (1.1) | 0 (00) |
| No | 68 (75.6) | 4.59±1.01 | 13 (14.4) | 45 (50.0) | 10 (11.1) |
| P value |  | <0.001 | <0.001<sup>a</sup> |  |  |
| Methods for adverse events |  |  |  |  |  |
| Yes | 36 (40.0) | 6.12±1.43 | 24 (26.7) | 12 (13.3) | 1 (1.1) |
| No | 54 (60) | 4.65±1.22 | 10 (11.1) | 34 (37.8) | 9 (10.0) |
| P value |  | <0.001 | <0.001<sup>a</sup> |  |  |

RCT: Randomized Controlled Trial; Student t-test; <sup>a</sup>chi-square test

## 4. DISCUSSION

In this review of methodological quality, COVID-19 clinical research articles were found to have a methodological flaw. Our findings support those of *Quinn TJ et*.*al*., who assessed the methodological and reporting quality of COVID-19 research papers published in major clinical journals (such as the BMJ, JAMA, The Lancet, and NEJM) from February to May 2020 and concluded that methodological and reporting issues could compromise the research’s utility. However, that study only examined the quality of scientific publications published in prominent clinical journals, which was a major limitation [9]. Our findings backed with the findings of a review of methodological quality of COVID-19 research articles published in February 2021 by Jung et al, concluded that the initial corpus of peer-reviewed COVID-19 research literature was mostly composed of observational studies that were submitted to a faster peer-review process and had lower methodological quality standards than comparable studies [10].

In our study, when comparing the modified Jadad scale score and the number of high-quality studies with the early and mid-late stages of publication, the modified Jadad scale score and the percentage of high-quality studies both increased. Early in the COVID-19 pandemic, an urgent demand for scientific data to support clinical, social, and economic decisions resulted in a shorter time to publication and an explosion of COVID-19 research published in both regular peer-reviewed journals and preprint servers. Furthermore, more than 30,000 of the COVID-19 articles published in 2020 were preprints, accounting for 17 to 30 percent of all COVID-19 research publications, and one-tenth of all preprints this year were about COVID-19, according to Dimensions [103]. This suggests that the expedited publishing of COVID-19 research, along with the accelerated review procedure, may have resulted in lower study quality scores.

We also investigated the impact of blinding and appropriate blinding in the COVID-19 RCTs, as well as their effects on study quality. We observed that there was a smaller number of the research reported with blinding and appropriate blinding, which significantly lowers the study quality. Blinding is a critical methodologic component of RCTs that helps to reduce bias and enhance the validity of the results. Since COVID-19 is a pandemic and RCT findings might impact clinical decision making, if participants are not blinded, their behavior in the trial and responses to subjective outcome measures may be influenced [104].

During the first wave of COVID-19, travel bans, quarantine, and stay-at-home measures were enacted, which were the principal barriers to doing efficient COVID-19-related research during the pandemic. The greatest concern during the clinical study was the threat of transmission of the COVID-19 infection not only to the participants but also to the researchers. Because of the small number of participants, the study had drawbacks such as a lack of power. To circumvent this hurdle, telemedicine, home medication delivery, web-based randomization, and online questionnaires were used to enroll participants in the trial [5]. However, findings of our results show that these methods resulted in the inadequacy of follow-up of participants, smaller sample size, and shorter study duration of the clinical trial, which may result in compromising the quality of the studies.

One of our study’s strengths is that we included all RCTs published on the prevention, diagnosis, treatment and management of COVID-19 from December 2019 to August 2021, and we used validated and standardized scales for quality evaluation, such as the modified Jadad scale.

Our study has certain drawbacks as well. Other quality evaluation criteria, such as sample size computation, sex reporting, and ethical approval, were not examined, and these scales can only assess methodology without regard for causal language.

In conclusion, we observed that the quality of RCTs published on COVID-19 during the pandemic has compromised. According to our findings, the expedited publishing of COVID-19 research, along with the faster review procedure, has resulted in lower study quality scores. Lack of quality can ultimately result in unreliable evidence, which will affect evidence-based clinical practice. So, there should be the adaptation of more reliable and cost-effective well-designed methods for conducting clinical trials without sacrificing data quality, ethics, safety, and efficacy and this should be a lesson learned from this catastrophe. With the emergence of stronger evidence, COVID-19 clinical studies with lower methodological quality should be revisited.

## Data Availability

All data produced in the present study are available upon reasonable request to the authors

## AUTHOR’S CONTRIBUTION

Conceptualization: KU, CT. Data curation: AJ, SS. Formal analysis: AJ, SS. Methodology: AJ, CT, SS, KU. Project administration: KU. Visualization – AJ, KU. Writing – original draft: AJ, CT. Writing – review & editing: AJ, CT, SS, KU.

