## supplementary appendix for "Can We Really Trust the Findings of the COVID-19 Research? Quality Assessment of Randomized Controlled Trials Published on COVID-19"

| ***Supplementary appendix S1: Search strategy*** | | | | |
| --- | --- | --- | --- | --- |
|  | **COVID-19 (#1)** | **Randomized controlled trial (#2)** | **Language (#3)** | **Date of publication (#4)** |
| **Search terms** | ("covid 19" OR "Coronavirus Infections” OR "severe acute respiratory syndrome coronavirus 2”) | “randomized controlled trial” OR "controlled clinical trial" OR “random” OR “randomly” OR “groups” OR “placebo” OR “trial” OR “ drug therapy” | “English” | "2019/12/01”: "2021/08/31" |
| **Search fields** | [Supplementary Concept], [MeSH Terms:noexp] | [Publication type], [Title/Abstract] , [MeSH SubHeading] | [Language] | [Date - Publication] |
| **Final search** | (#1 AND #2 AND #3 AND #4) ^*^ | | | |

***Supplementary appendix S2: modified Jadad scale***

| **Item** | **Points** | **Description** |
| --- | --- | --- |
| Randomization | 2 | 1 point if randomization is mentioned.  1 additional point if the method of randomization is appropriate. (A method to generate the sequence of randomization will be regarded as appropriate if it allowed each study participant to have the same chance of receiving each intervention and the investigators could not predict which treatment was next. Methods of allocation using the date of birth, date of admission, hospital numbers, or alternation should not be regarded as appropriate)  1 point will be deducted if the method of randomization is inappropriate. (Patients were allocated alternately, or according to date of birth, hospital number, etc.) |
| Blinding | 2 | 1 point if a double binding is described.  1 additional point if the method of double-blinding is appropriate. (A study must be regarded as double-blind if the word “double-blind” is used. The method will be regarded as appropriate if it is stated that neither the person doing the assessments nor the study participant could identify the intervention being assessed, or if in the absence of such a statement the use of active placebos, identical placebos, or dummies is mentioned.)  1 point will be deducted if blinding was inappropriate (e.g., comparison of tablet vs. injection with no double-dummy) |
| Withdrawals or dropouts | 1 | Participants who were included in the study but did not complete the observation period or who were not included in the analysis must be described. The number and the reasons for withdrawal in each group must be stated. If there were no withdrawals, it should be stated in the article. If there is no statement on withdrawals, this item must be given no points. |
| Method to assess adverse events | 1 | If not described, this item must be given no points. |
| Statistical analysis | 1 | If not described, this item must be given no points. |
| Inclusion or exclusion criteria | 1 | If not described, this item must be given no points. |
